# AIDCOV: An Interpretable Artificial Intelligence Model for Detection of COVID-19 from Chest Radiography Images

**DOI:** 10.1101/2020.05.24.20111922

**Authors:** Maryam Zokaeinikoo, Pooyan Kazemian, Prasenjit Mitra, Soundar Kumara

## Abstract

As the Coronavirus Disease 2019 (COVID-19) pandemic continues to grow globally, testing to detect COVID-19 and isolating individuals who test positive remains to be the primary strategy for preventing community spread of the disease. The current gold standard method of testing for COVID-19 is the reverse transcription polymerase chain reaction (RT-PCR) test. The RT-PCR test, however, has an imperfect sensitivity (around 70%), is time-consuming and labor-intensive, and is in short supply, particularly in resource-limited countries. Therefore, automatic and accurate detection of COVID-19 using medical imaging modalities such as chest X-ray and Computed Tomography, which are more widely available and accessible, can be beneficial as an alternative diagnostic tool. We develop a novel hierarchical attention neural network model to classify chest radiography images as belonging to a person with either COVID-19, other infections, or no pneumonia (i.e., normal). We refer to this model as Artificial Intelligence for Detection of COVID-19 (AIDCOV). The hierarchical structure in AIDCOV captures the dependency of features and improves model performance while the attention mechanism makes the model interpretable and transparent. Using a publicly available dataset of 5801 chest images, we demonstrate that our model achieves a mean cross-validation accuracy of 97.8%. AIDCOV has a sensitivity of 99.3%, a specificity of 99.98%, and a positive predictive value of 99.6% in detecting COVID-19 from chest radiography images. AIDCOV can be used in conjunction with or instead of RT-PCR testing (where RT-PCR testing is unavailable) to detect and isolate individuals with COVID-19 and prevent onward transmission to the general population and healthcare workers.

## Introduction

The outbreak of the novel coronavirus known as Severe Acute Respiratory Syndrome CoronaVirus 2 (SARS-CoV-2) started in Wuhan, China, in December 2019 and rapidly spread worldwide. As the number of people with Coronavirus Disease 2019 (COVID-19) escalates in the United States and around the world, reducing the number of transmissions from infected individuals to the general population and healthcare workers becomes increasingly important and challenging.

Although about 8 in 10 people who contract the SARS-CoV-2 virus remain asymptomatic or develop only mild to moderate symptoms (Mizumoto et al. 2020; Arons et al. 2020; Wu and McGoogan 2020; Yang et al. 2020), others may develop life-threatening conditions, such as dyspnea, pneumonia, or severe acute respiratory syndrome, which require hospital or ICU care with supplemental oxygen or mechanical ventilation (Mahase 2020). The rapid spread of COVID-19 is, in part, due to the lack of sufficient testing and isolation of positive cases, which subsequently leads to community transmission from undiagnosed cases. This rapid spread may result in overwhelming and collapsing healthcare systems, even in developed countries, due to the surge in demand for hospital and ICU care.

A critical step in controlling the transmissions and flattening the curve is to use a widely-available, fast, and accurate COVID-19 detection method, and to immediately isolate diagnosed cases until they are no longer infectious.

The current gold standard screening method for COVID-19 is the direct detection of SARS-CoV-2 RNA by reverse transcription polymerase chain reaction (RT-PCR) test (Patel, Jernigan, and others 2020). Several RT-PCR assays are used in the U.S. and around the world. Each has different performance characteristics and turnaround time (ranging from minutes to several hours) and requires different specimen types (World Health Organization 2020). The sensitivity of RT-PCR testing is widely variable; depending on the assay, the type and quality of the specimen obtained, the stage of the disease, and the duration of infection, it can vary between 32% to 73% (Wang et al. 2020a; Kucirka et al. 2020; Guo et al. 2020). Therefore, there is an immediate need for accessible, rapid, and accurate testing tools to help combat the spread of the SARS-CoV-2 virus.

Medical imaging modalities such as Chest X-Ray (CXR) and Computed Tomography (CT) can be used as an alternative to RT-PCR testing to detect characteristic symptoms of COVID-19 in patients’ chest images (Xu et al. 2020; Ng et al. 2020). Detecting COVID-19 from chest radiography images has shown promising results and higher sensitivity compared to RT-PCR testing (Fang et al. 2020). Moreover, CT images of patients with COVID-19 may show abnormalities before the patient develops symptoms and before the detection of viral RNA from upper respiratory specimens (Sutton et al. 2020; Zhao et al. 2020). However, visual evaluation of radiography images to find subtle signs of COVID-19 is both fallible and time-consuming. In this context, artificial intelligence (AI) methods can be leveraged to analyze medical images for subtle signs of SARS-CoV-2 infection automatically and to detect COVID-19 rapidly and accurately.

In this study, we develop a new deep learning model for detection of COVID-19 using CXR and CT scan images automatically. We refer to this model as Artificial Intelligence for Detection of COVID-19 (AIDCOV). AIDCOV employs a novel hierarchical attention structure, which can tell clinicians the specific locations of the lung affected by the SARS-CoV-2 infection rapidly and with high sensitivity and specificity.

## Methods

AIDCOV includes a novel two-level hierarchical attention structure for classification of chest radiography images into one of the three classes: COVID-19 viral infection, other viral/bacterial infection (i.e., non-COVID-19 infection), or normal (i.e., no infection). This hierarchical structure enables the model to capture the dependency of features extracted from chest images via a pre-trained network (e.g., VGG-16) in both horizontal and vertical directions and helps improve model performance. The attention mechanism makes the black-box deep neural network model interpretable such that the model can designate the specific locations of patients’ lungs that manifest subtle signs of infection. AIDCOV is an end-to-end deep neural network model, which does not require any feature engineering.

### Transfer Learning

Deep neural network models often include hundreds of thousands of hyperparameters, and thus they need to be trained on very large datasets. Transfer learning is a technique that allows training deep neural network models on small datasets by taking a pre-trained deep neural network model and repurposing it for a different task. We leverage the powerful idea of transfer learning by employing VGG-16 (Simonyan and Zisserman 2014), a pre-trained convolutional neural network that is trained on a dataset of more than 15 million images (Krizhevsky, Sutskever, and Hinton 2012). VGG-16 has shown promising performance for medical image analysis (Yadav and Jadhav 2019; Shen et al. 2019; Guan et al. 2019). VGG-16 has 13 convolutional layers, 5 pooling layers, and 3 fully connected layers. We removed the 3 fully connected layers and replaced them with our novel hierarchical attention structure. While the early layers of VGG-16 learn low-level features of the image, our hierarchical attention model learns subtle signs of COVID-19 and other viral/bacterial infections and determines the final classification.

### Two-level Hierarchical Deep Neural Network Model for Image Classification

In this section, we describe our novel hierarchical attention structure for image classification, which considers the dependencies of feature components in both horizontal (width) and vertical (height) directions.

#### Step 1: Resizing the image

First, we need to resize input chest radiography images to the format that is compatible with the pre-trained VGG-16 model. We resized all the input images to size 160 × 160 × 3.

#### Step 2: Feature extraction

We use VGG-16 to obtain a low-dimensional feature representation vector for each resized image (Figure 1).

**Figure 1:**
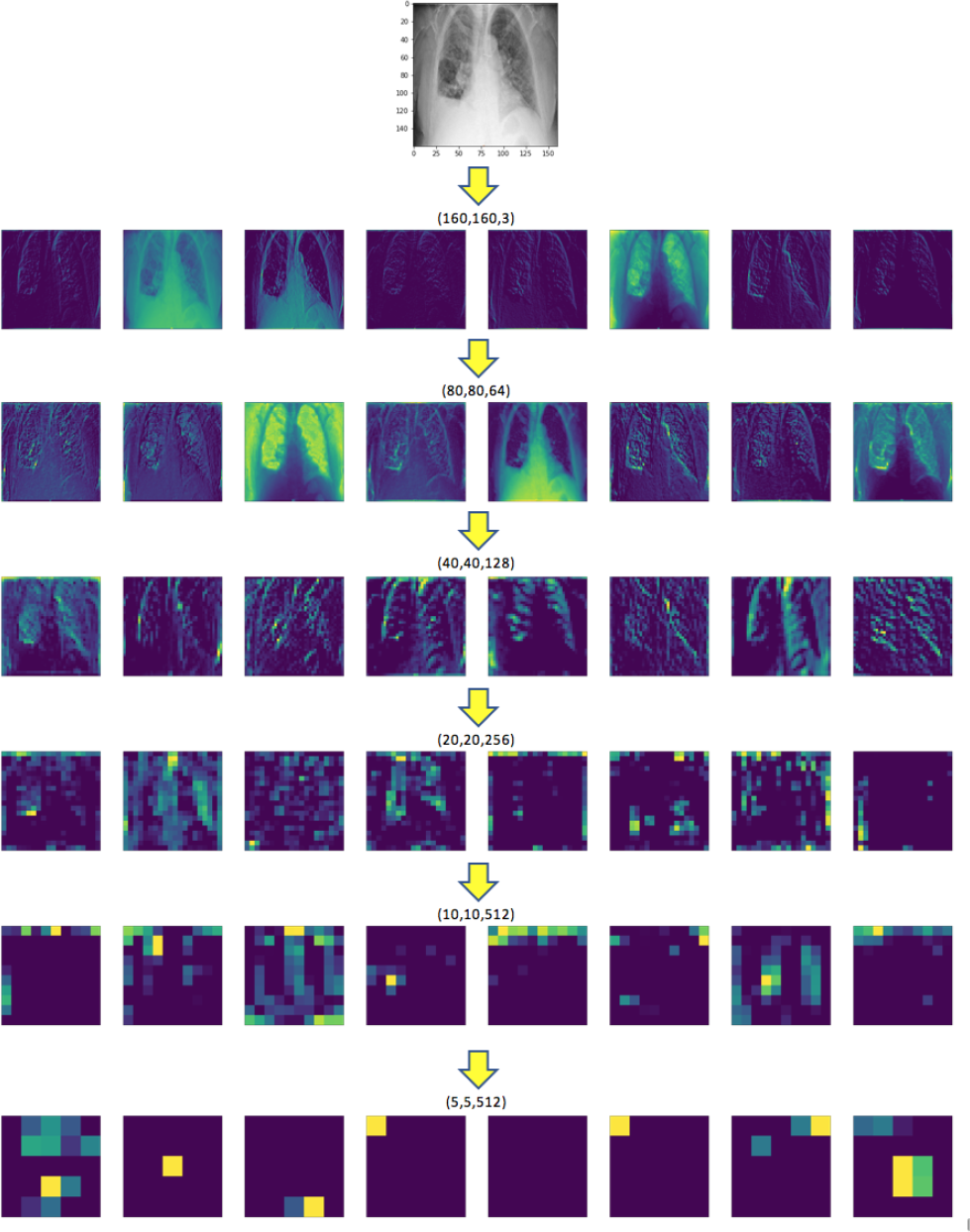
Low-dimensional feature extraction using the pretrained VGG-16 model.

In general, if the input image is of size (A, B, 3), the output of the VGG-16 model will be a tensor of size (A/32,B/32, 512). In our case, since the input image is of size (160,160,3), the output of the model is of size (5, 5, 512). We show this output by *X*, which is obtained as follows:

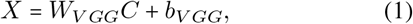

where *C* is the resized radiography image, and *W*_*V GG*_ and *b*_*V GG*_ are the trained weight and bias matrices obtained from the pre-trained VGG-16 model.

As mentioned above, *X* is the output of size (5, 5, 512). We refer to each (1, 1, 512) block of *X* as *x*_*ij*_, where *i* ∈ [1, 5] and *j* ∈ [1, 5] (Figure 2).

**Figure 2:**
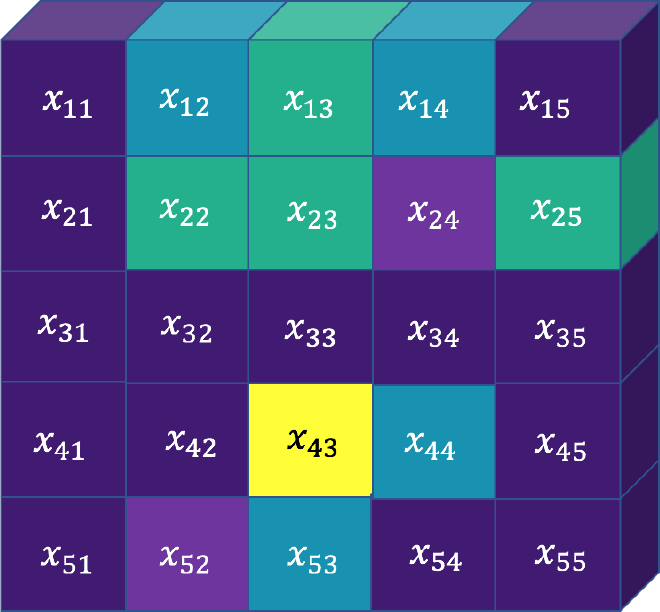
The output block of size 5 × 5 × 512.

#### Step 3: Horizontal feature encoding

Next, for each level of *i* ∈ [1, 5], we are going to encode the output block along the *x* axis (i.e., horizontally). To do so, we apply a GRU-BRNN on *x*_*i*1_, …, *x*_*i*5_ for each level of *i* to incorporate horizontal dependencies (in both forward and backward directions) within each row of an image. We have,

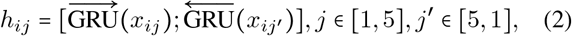

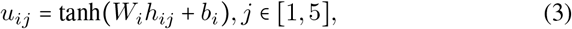

where *h*_*ij*_ is the output of the GRU-BRNN and *u*_*ij*_ is its hidden representation. *W*_*i*_ and *b*_*i*_ are the weight matix and bias vector for each row of the *x*_*ij*_ block learned through training. Since we aim to determine the contribution of each *x*_*ij*_ block within each row (horizontal level) to the overall prediction, we applied an attention layer on top of the hidden representations *u*_*ij*_ to obtain the attention scores *α*_*ij*_ by learning the row context vector *u*_*i*_,

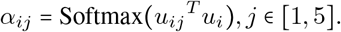

Finally, we encode each *ŷ*_*i*_ block as a weighted sum of *h*_*ij*_ and the attention scores *α*_*ij*_ (Figure 3),

**Figure 3:**
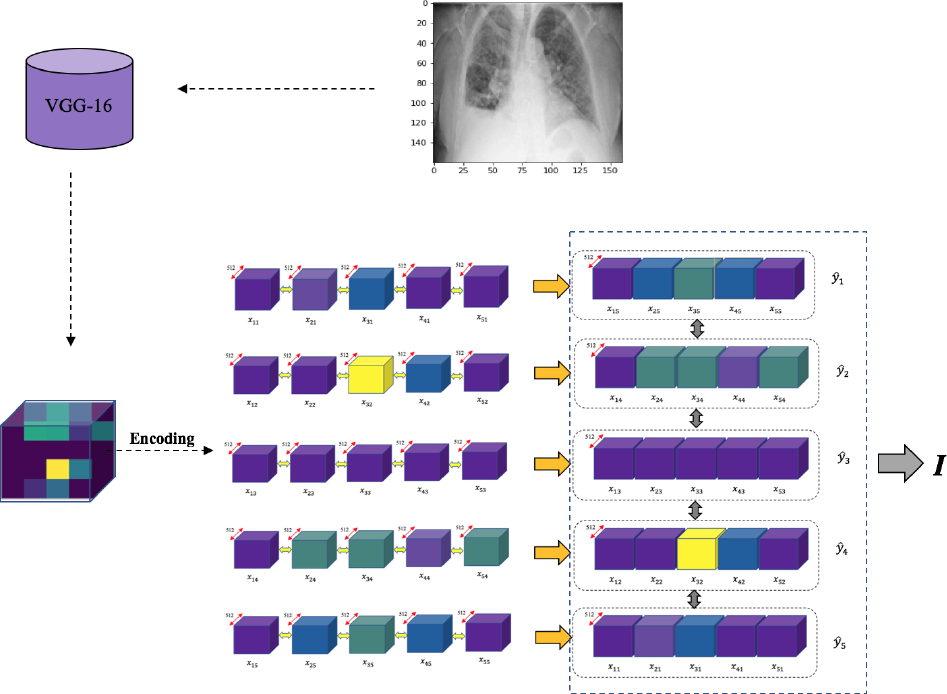
Encoding feature outputs in both *x* and *y* axis.

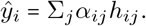

#### Step 4: Vertical feature encoding

In this step, we encode the representations *ŷ*_*i*_ computed from the previous step along the *y* axis (i.e., vertically). We further determine the attention scores *α*_*i*_ for each *ŷ*_*i*_ block as follows.

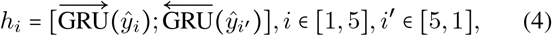

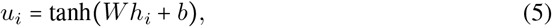

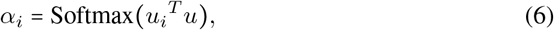

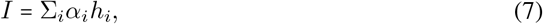

where *h*_*i*_ capture the dependencies of *ŷ*_*i*_ blocks using GRU-BRNN and *u*_*i*_ is its hidden representation obtained through training of *W* and *b* parameters. The attention weights *α*_*i*_ for *ŷ*_*i*_ are computed using *u*_*i*_ and the trained context vector of *u*. The image encoding is the weighted sum of *h*_*i*_ encodings and attention scores *α*_*i*_. Finally, we use the image encoding *I* to build a multi-class classifier as,

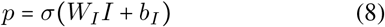

Figure 4 shows the hierarchical encoding network of Steps 3 and 4 described above.

**Figure 4:**
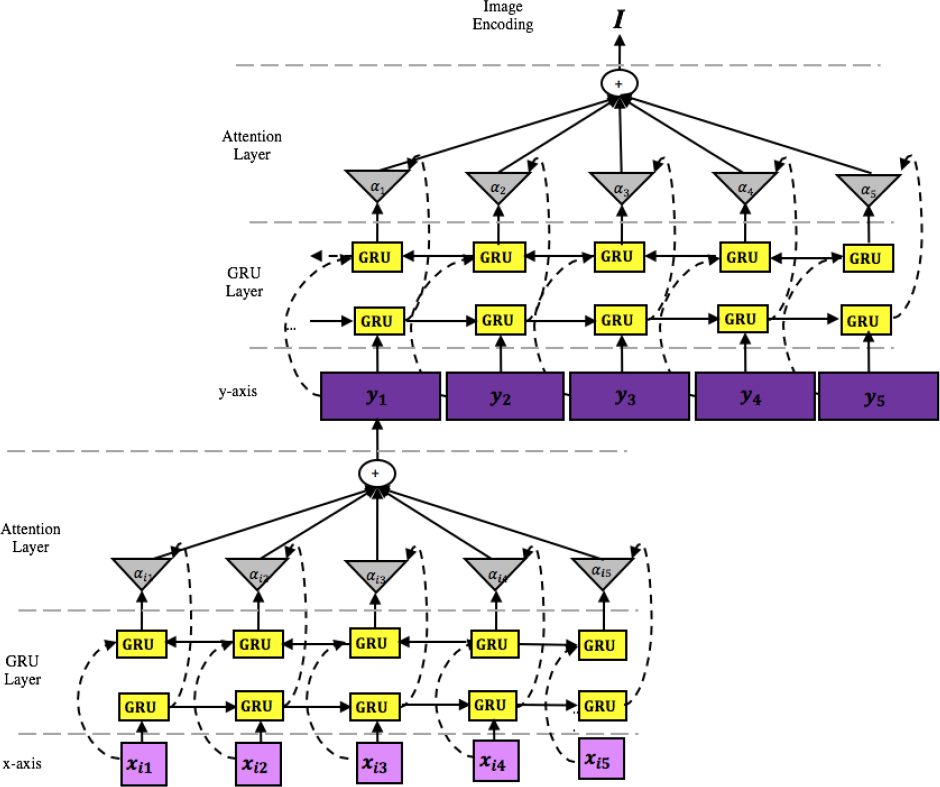
The hierarchical attention structure for image encoding.

### Data

We leverage a labeled dataset of 5801 chest X-ray images and CT scans to train and test the AIDCOV model. This dataset consists of 269 images from patients with COVID-19 obtained in early April 2020 from an open-source GitHub repository (Cohen, Morrison, and Dao 2020), which can be accessed on the web: https://github.com/ieee8023/covid-chestxray-dataset. Moreover, our dataset includes 3949 images from patients with other viral/bacterial infections as well as 1583 images from individuals with no pneumonia (i.e., normal), both obtained from the “Chest X-Ray Images” Kaggle repository (Kaggle 2013). Therefore, each image in our dataset is labeled as either COVID-19, other infections, or normal. This study was exempt from institutional review board (IRB) review since it used publicly available, deidentified data.

### Implementation Details

We set the GRU dimension to 50 for both horizontal (Step 3) and vertical (Step 4) encoding levels. So, the bidirectional GRU has 100 dimensions. We used a mini-batch size of 20 images and trained on 10 epochs.

We used ‘categorical cross entropy’ as our loss function to classify chest radiography images into one of the three classes (COVID-19, other infection, or normal). We employed the Adaptive Subgradient (Adagrad) as the optimizer.

To evaluate the model’s performance, we conducted 10-fold stratified cross-validation such that in each fold, we trained our model on 4698 samples, validated on 523 samples, and tested it on 580 samples. Each set of stratified cross-validation includes the same ratio of subjects of each class in each fold.

### Comparison with Simpler Structures

To better evaluate the value of our novel hierarchical attention structure, we developed two related but simpler deep learning models. In one model, we removed the attention mechanism from our base model, i.e., we fed the feature representations obtained from VGG-16 to the hierarchical structure of Figure 4 without the attention layers. In another model, we replaced the whole hierarchical attention network with a fully connected network.

## Results

### Model Performance

AIDCOV achieved a mean cross-validation accuracy of 97.8% across the 10 folds (Table 1). The model demonstrated excellent performance in detecting COVID-19. The hierarchical attention model had a sensitivity (true positive rate) of 99.3%, a specificity (true negative rate) of 99.98%, and a positive predictive value (PPV) of 99.6% for detecting COVID-19 from chest radiography images (Figure 2).

**Table 1:** Results: 10-fold stratified cross validation

| Fold | Accuracy | Normal | COVID-19 | Non-COVID19 |
| --- | --- | --- | --- | --- |
| 1 | 97.6 | 96.9 | 96.0 | 98.0 |
| 2 | 96.7 | 93.9 | 96.0 | 97.8 |
| 3 | 96.2 | 87.8 | 100 | 99.5 |
| 4 | 97.2 | 94.7 | 100 | 98.2 |
| 5 | 97.8 | 96.4 | 100 | 98.2 |
| 6 | 98.8 | 98.2 | 100 | 98.9 |
| 7 | 98.1 | 96.2 | 100 | 98.7 |
| 8 | 98.8 | 95.9 | 100 | 99.8 |
| 9 | 98.8 | 97.2 | 100 | 99.3 |
| 10 | 98.4 | 96.8 | 100 | 99.0 |
| Mean | 97.8 | 95.4 | 99.2 | 98.7 |

**Table 2:** Confusion matrix

|  |  | Prediction |  |  |
| --- | --- | --- | --- | --- |
|  |  | Normal | COVID-19 | Other Infection |
| Ground Truth | Normal | 1510 | 0 | 73 |
|  | COVID-19 | 1 | 267 | 1 |
|  | Other Infection | 49 | 1 | 3899 |

AIDCOV also demonstrated promising performance for correctly classifying non-COVID-19 chest images. The model had a sensitivity (specificity) of 98.7% (96.0%) for detecting other viral/bacterial infections and a sensitivity (specificity) of 95.4% (98.8%) for normal chest radiography images. The PPV for other infections and normal images were 98.1% and 96.8%, respectively (Figure 2). These results suggest that AIDCOV performs well in detecting COVID-19, other viral/bacterial infections, and normal cases based on the chest radiography images.

### The Value of Hierarchical Attention Structure

The hierarchical model (without the attention layers) and the fully connected structure had lower accuracy levels than the hierarchical attention model. The hierarchical (no attention) model had an overall cross-validation accuracy of 97.5%, slightly lower than the hierarchical attention model. The sensitivity of this model to detect COVID-19 from chest radiography images was 99.3%, similar to the hierarchical attention model. The model with a fully connected structure had the poorest performance among the three models and resulted in an overall cross-validation accuracy of 96.0% when tested on our dataset. This model had a sensitivity of 93.3% in detecting COVID-19.

These results highlight the value of our novel hierarchical structure, which can capture the dependency of all feature representation blocks (obtained from the VGG-16) in both horizontal and vertical directions and improve model performance. The attention mechanism in our hierarchical attention model also helps with the interpretability and transparency of the model predictions.

### Interpretability

The strengths of AIDCOV are not limited to its superior sensitivity, specificity, and PPV in detecting COVID-19 and other infections. To gain deeper insights into how the model makes its predictions and identify the areas of the lung affected by the infection, we extracted attention scores for each image. Figure 5 illustrates the areas of the chest radiography images for each type (COVID-19, other infections, and normal) that the model paid more attention to based on the final attention scores averaged over all images of that type. It can be seen that the model identified signs of SARS-CoV-2 infection mostly in the lower zone and other infections around the middle zone of the lung. Since the model determines the attention scores relatively, the normal radiography images had the highest attention score on the very top level corresponding to the upper zone of the lung. In other words, since the lower and middle zones of the lung contain signs of COVID-19 and other infections and, therefore, dominate the attention scores for these zones, the normal images receive lower attention scores for the middle and lower zones and higher attention scores for the upper zone of the lung.

**Figure 5:**
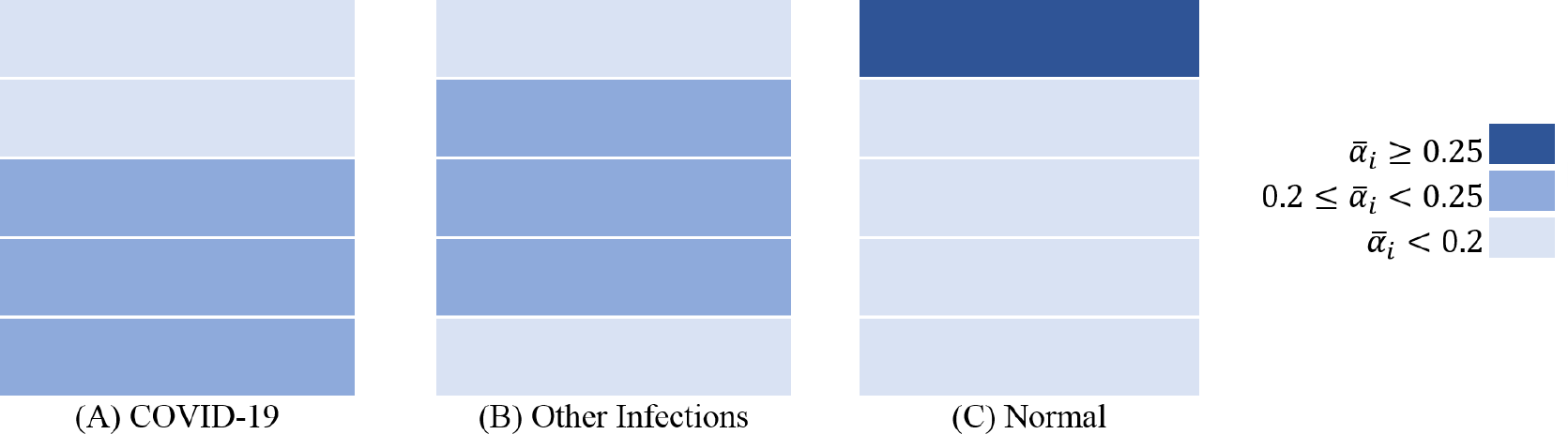
Average attention score for different zones of the lung for each image class.

Moreover, AIDCOV can identify the specific blocks within each individual’s chest image that may include subtle signs of infection via the attention scores of both encoding levels (i.e., horizontal and vertical). To better illustrate the model’s interpretability, we included the chest images of three patients with COVID-19 in Figure 6 and provided attention scores for each zone of the lung and each block of the image. Given that there are five zones in each image, we highlighted the zones that received an attention score of 0.2 or higher. Then, within those zones, we highlighted the blocks that received an attention score greater than or equal to 0.2.

**Figure 6:**
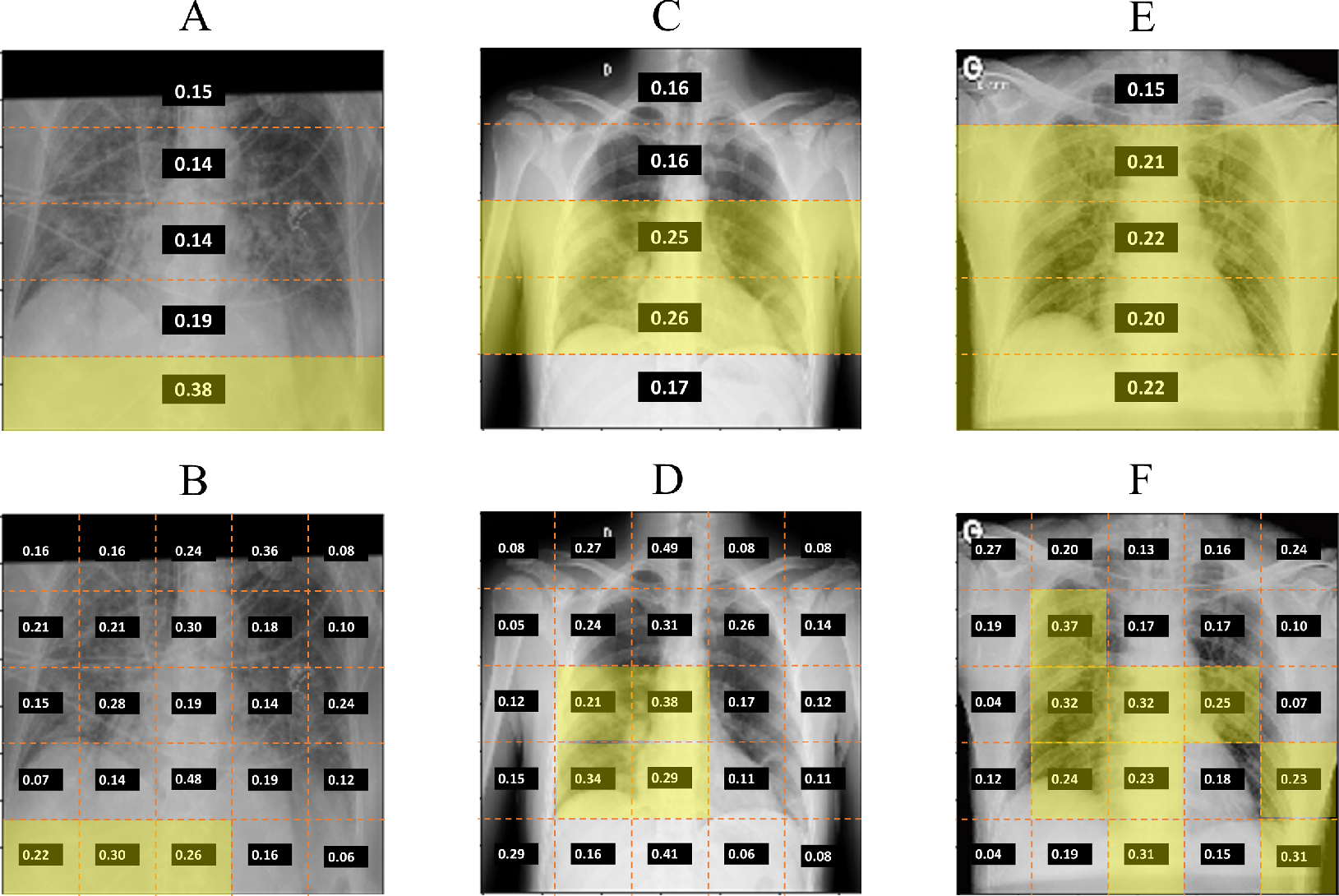
Attention scores for different zones of the lung (horizontal level) and different blocks of the image for 3 patients with COVID-19. Signs of COVID-19 were detected in the lower zone for Patient 1 (A-B), middle zone for Patient 2 (C-D), and lower and middle zones for Patient 3 (E-F). about 10% of infections were among the healthcare workers (Wang, Zhou, and Liu 2020). Thus, leveraging chest radiographs in hospitals to identify patients with COVID-19 and using appropriate personal protective equipment (PPE) when providing care to these patients can be beneficial.

In Figure 6, the first patient (Panels A and B) was admitted to the hospital with fever, shortness of breath (dyspnea), and low oxygen saturation. Radiological worsening with changes within the lower lobes was noted on her chest Xray. Our model correctly gave a higher attention score to the lower zone of the lung and identified the specific areas of the lower zone that demonstrated signs of SARS-CoV-2 infection.

The second patient (Panels C and D) in Figure 6 came to the hospital with suspected pneumonia. The radiographic investigation indicated abnormalities in the middle part of the right lung. As seen in Figure 6.C and 6.D, our model correctly identified these abnormalities in the middle zone of the right lung. (Note that the right lung appears on the left side of the radiography image.)

The chest image of the third patient (Panels E and F) in Figure 6 indicated small consolidation in the right upper lobe and ground-glass opacities in both lower lobes. Our deep learning model correctly identified the middle and the lower zones as the areas with abnormalities (Figure 6.E). It further pointed to the right lung (that appears on the left side of Figure 6.F) and the lower zone of both lungs.

## Discussion

In this study, we introduced AIDCOV, an artificial intelligence model for detection of COVID-19 from chest radiography images. The model performs multi-class prediction, i.e., it labels each image as belonging to a person with either COVID-19, other infections, or no pneumonia (i.e., normal). AIDCOV leverages VGG-16 to obtain a low-dimensional feature representation for each image. It then encodes the features along the horizontal (width) and vertical (height) directions using a novel two-level hierarchical attention structure. This allows the model to capture the horizontal and vertical dependencies of the features, which is ignored in a fully connected network. The attention mechanism further helps make the model interpretable and gives transparency to model predictions.

We trained and tested AIDCOV on publicly available datasets comprising 5801 chest radiography images. Of these, 269 samples had COVID-19, 3949 samples had other viral/bacterial infections, and the remaining 1583 samples were normal. We demonstrated that the model has an overall accuracy of 97.8% across the ten folds of cross-validation. AIDCOV showed excellent sensitivity (99.3%), specificity (99.98%), and positive predictive value (99.6%), in detecting COVID-19 from chest radiography images.

The high sensitivity and specificity of our model are critical in practice since a false negative result can lead to not isolating an individual with COVID-19, which can subsequently result in many transmissions from that person to others, including to the healthcare workers. Transmission from patients to healthcare workers results in undermining the healthcare capacity. In China, about 5%, and in Italy,

The high positive predictive value of AIDCOV implies a very low rate of false positives. This is also crucial because false-positive results can put additional burden on the healthcare system due to unnecessary use of scarce resources such as hospital isolation rooms (e.g., negative pressure rooms) and PPE for healthcare workers, which could be used for actual COVID-19 patients. These results indicate that the AIDCOV model can be a reliable tool for detecting COVID-19 from chest radiography images.

To better assess the value of our hierarchical attention structure, we developed two simpler models. In one model, we kept the hierarchical structure but removed the attention mechanism from it. In another model, we replaced the whole hierarchical attention structure with a fully connected network. The model with a fully connected network had the poorest performance among the three models and achieved an accuracy of 96.0%. The hierarchical model without attention had an accuracy of 97.5%, which is only slightly below our hierarchical attention model’s accuracy. However, the main advantage of including an attention mechanism is making the model interpretable and providing transparency to model predictions.

Using an analysis of the attention scores that AIDCOV gave to COVID-19, other infections, and normal samples, we demonstrated that the model identified signs of SARS-CoV-2 infection, on average, in the lower and middle zones of the lung more frequently. This is consistent with findings from radiology reports, which indicate that abnormalities due to SARS-CoV-2 infection are more commonly found in inferior and middle lobes of the lung, corresponding to the lower and middle zones of the chest radiography images (Pan et al. 2020; Zhou et al. 2020; Bernheim et al. 2020; Zu et al. 2020; Wong et al. 2020). Furthermore, our hierarchical attention model divides each image into 25 blocks (5 × 5) and provides the attention score for each block. This can shed light on particular regions within each radiography image that may contain subtle signs of infection and makes the model more transparent and trustable.

Currently, the primary screening tool to detect COVID-19 is reverse transcription polymerase chain reaction (RT-PCR), which is a laboratory test to detect viral nucleic acid (Patel, Jernigan, and others 2020). However, not only the capacity for RT-PCR testing is limited (both in the U.S. and in many other countries), but also the result return time can range between several minutes to hours (World Health Organization 2020). More importantly, the RT-PCT test has a sensitivity of around 70%, which can be even lower depending on the assay, type and quality of the specimen, and the disease stage (Huang et al. 2020; Ai et al. 2020; Wang et al. 2020a; Kucirka et al. 2020; Guo et al. 2020). This means that about three in ten individuals with COVID-19 receive a false-negative test result. In this context, AIDCOV provides an accurate and fast alternative to RT-PCR testing that can quickly detect COVID-19 from chest radiography images. Moreover, given that RT-PCR testing kits are in short supply in many resource-limited settings, using chest Xray and CT scan images, which are generally more available around the world, as a screening tool for COVID-19 may be worth considering. We demonstrated that our model is a viable alternative to RT-PCR testing, which can be used to detect and quarantine infected individuals and stop the spread of the SARS-CoV-2 virus.

A number of other artificial intelligence models to detect COVID-19 from chest radiography images have also been developed recently. Li et al. developed COVNet, a neural network model to detect COVID-19 and community-acquired pneumonia from CT images (Li et al. 2020). COVNet demonstrated a sensitivity of 89.8% and a specificity of 95.8% in detecting COVID-19. It also achieved a sensitivity and specificity of 86.9% and 92.3%, respectively, in detecting community-acquired pneumonia from CT images. Wang et al. developed COVID-Net using a deep convolutional neural network structure designed for detecting COVID-19 and other infections from chest X-ray images (Wang, Lin, and Wong 2020). COVID-Net reports a sensitivity of 91.0% and a positive predictive value (PPV) of 98.9% in detecting COVID-19. The sensitivity and PPV of COVID-Net in detecting other infections were 94.0% and 91.3%, respectively. Zhang et al. developed a deep learning model to detect COVID-19 from chest X-ray images (Zhang et al. 2020). Their model had a sensitivity (specificity) of 72.0% (98.0%) with a threshold of 0.5 and 96.0% (70.7%) with a threshold of 0.15. Other studies evaluated a number of convolutional neural network structures on very small datasets of chest radiography images and achieved various accuracy levels (Hemdan, Shouman, and Karar 2020; Narin, Kaya, and Pamuk 2020). AIDCOV outperforms the models described above in terms of sensitivity, specificity, and PPV in detecting COVID-19 and other infections from chest radiography images.

Our study has a number of limitations and, therefore, our results should be interpreted with caution. First, as was the case with the other related studies, our dataset was limited in size and had only 269 cases with COVID-19. Further validation on datasets with a larger number of chest radiography images from patients with COVID-19 would be valuable. Second, chest Xray images may not show signs of SARS-CoV-2 infections in the early stages of illness. Abnormalities are more likely to develop over the course of the disease (Wang et al. 2020b; Simpson et al. 2020). However, some preliminary data suggest that abnormalities may show in CT images in the presymptomatic stage and prior to the detection of viral RNA from upper respiratory specimens (Sutton et al. 2020; Zhao et al. 2020). Our dataset does not include information on time since symptom onset or the disease stage at the time the image was taken; thus, we could not assess our model’s accuracy based on these factors.

In conclusion, AIDCOV demonstrated high sensitivity, specificity, and positive predictive value in detecting COVID-19 from chest Xray and CT images. Given that radiography is widely available in many countries around the world, AIDCOV can be used in conjunction with or instead of RT-PCR testing (e.g., where RT-PCR testing is unavailable) to find individuals infected with the SARS-CoV-2 virus, isolate them, and prevent the spread of COVID-19.

## Data Availability

We used publicly available data sources. The links are included in the manuscript.

## References

Ai, T.; Yang, Z.; Hou, H.; Zhan, C.; Chen, C.; Lv, W.; Tao, Q.; Sun, Z.; and Xia, L. 2020. Correlation of chest ct and rtpcr testing in coronavirus disease 2019 (covid-19) in china: a report of 1014 cases. Radiology 200642.

Arons, M. M.; Hatfield, K. M.; Reddy, S. C.; Kimball, A.; James, A.; Jacobs, J. R.; Taylor, J.; Spicer, K.; Bardossy, A. C.; Oakley, L. P.; et al. 2020. Presymptomatic sars-cov-2 infections and transmission in a skilled nursing facility. New England Journal of Medicine.

Bernheim, A.; Mei, X.; Huang, M.; Yang, Y.; Fayad, Z. A.; Zhang, N.; Diao, K.; Lin, B.; Zhu, X.; Li, K.; et al. 2020. Chest ct findings in coronavirus disease-19 (covid-19): relationship to duration of infection. Radiology 200463.

Cohen, J. P.; Morrison, P.; and Dao, L. 2020. Covid-19 image data collection. 2003.11597.

Fang, Y.; Zhang, H.; Xie, J.; Lin, M.; Ying, L.; Pang, P.; and Ji, W. 2020. Sensitivity of chest ct for covid-19: comparison to rt-pcr. Radiology 200432.

Guan, Q.; Wang, Y.; Ping, B.; Li, D.; Du, J.; Qin, Y.; Lu, H.; Wan, X.; and Xiang, J. 2019. Deep convolutional neural network vgg-16 model for differential diagnosing of papillary thyroid carcinomas in cytological images: a pilot study. Journal of Cancer 10(20):4876.

Guo, L.; Ren, L.; Yang, S.; Xiao, M.; Chang, D.; Yang, F.; Dela Cruz, C. S., Wang, Y.; Wu, C.; Xiao, Y.; et al. 2020. Profiling early humoral response to diagnose novel coronavirus disease (covid-19). Clinical Infectious Diseases.

Hemdan, E. E.-D.; Shouman, M. A.; and Karar, M. E. 2020. Covidx-net: A framework of deep learning classifiers to diagnose covid-19 in x-ray images. arXiv preprint 2003.11055.

Huang, P.; Liu, T.; Huang, L.; Liu, H.; Lei, M.; Xu, W.; Hu, X.; Chen, J.; and Liu, B. 2020. Use of chest ct in combination with negative rt-pcr assay for the 2019 novel coronavirus but high clinical suspicion. Radiology 295(1):22–23.

Kaggle. 2013. Chest x-ray images (pneumonia). https://www.kaggle.com/paultimothymooney/chest-xray-pneumonia, Accessed on May 24, 2020.

Krizhevsky, A.; Sutskever, I.; and Hinton, G. E. 2012. Imagenet classification with deep convolutional neural networks. In Advances in neural information processing systems, 1097–1105.

Kucirka, L. M.; Lauer, S. A.; Laeyendecker, O.; Boon, D.; and Lessler, J. 2020. Variation in false-negative rate of reverse transcriptase polymerase chain reaction–based sars-cov-2 tests by time since exposure. Annals of Internal Medicine.

Li, L.; Qin, L.; Xu, Z.; Yin, Y.; Wang, X.; Kong, B.; Bai, J.; Lu, Y.; Fang, Z.; Song, Q.; et al. 2020. Artificial intelligence distinguishes covid-19 from community acquired pneumonia on chest ct. Radiology 200905.

Mahase, E. 2020. Coronavirus: covid-19 has killed more people than sars and mers combined, despite lower case fatality rate.

Mizumoto, K.; Kagaya, K.; Zarebski, A.; and Chowell, G. 2020. Estimating the asymptomatic proportion of coronavirus disease 2019 (covid-19) cases on board the diamond princess cruise ship, yokohama, japan, 2020. Eurosurveillance 25(10):2000180.

Narin, A.; Kaya, C.; and Pamuk, Z. 2020. Automatic detection of coronavirus disease (covid-19) using x-ray images and deep convolutional neural networks. arXiv preprint 2003.10849.

Ng, M.-Y.; Lee, E. Y.; Yang, J.; Yang, F.; Li, X.; Wang, H.; Lui, M. M.-s.; Lo, C. S.-Y.; Leung, B.; Khong, P.-L.; et al. 2020. Imaging profile of the covid-19 infection: radiologic findings and literature review. Radiology: Cardiothoracic Imaging 2(1):e200034.

Pan, F.; Ye, T.; Sun, P.; Gui, S.; Liang, B.; Li, L.; Zheng, D.; Wang, J.; Hesketh, R. L.; Yang, L.; et al. 2020. Time course of lung changes on chest ct during recovery from 2019 novel coronavirus (covid-19) pneumonia. Radiology 200370.

Patel, A.; Jernigan, D. B.; et al. 2020. Initial public health response and interim clinical guidance for the 2019 novel coronavirus outbreak—united states, december 31, 2019– february 4, 2020. Morbidity and Mortality Weekly Report 69(5):140.

Shen, L.; Margolies, L. R.; Rothstein, J. H.; Fluder, E.; McBride, R.; and Sieh, W. 2019. Deep learning to improve breast cancer detection on screening mammography. Scientific reports 9(1):1–12.

Simonyan, K.; and Zisserman, A. 2014. Very deep convolutional networks for large-scale image recognition. arXiv preprint 1409.1556.

Simpson, S.; Kay, F. U.; Abbara, S.; Bhalla, S.; Chung, J. H.; Chung, M.; Henry, T. S.; Kanne, J. P.; Kligerman, S.; Ko, J. P.; et al. 2020. Radiological society of north america expert consensus statement on reporting chest ct findings related to covid-19. endorsed by the society of thoracic radiology, the american college of radiology, and rsna. Radiology: Cardiothoracic Imaging 2(2):e200152.

Sutton, D.; Fuchs, K.; D’alton, M.; and Goffman, D. 2020. Universal screening for sars-cov-2 in women admitted for delivery. New England Journal of Medicine.

Wang, W.; Xu, Y.; Gao, R.; Lu, R.; Han, K.; Wu, G.; and Tan, W. 2020a. Detection of sars-cov-2 in different types of clinical specimens. Jama 323(18):1843–1844.

Wang, Y.; Liu, Y.; Liu, L.; Wang, X.; Luo, N.; and Li, L. 2020b. Clinical outcomes in 55 patients with severe acute respiratory syndrome coronavirus 2 who were asymptomatic at hospital admission in shenzhen, china. The Journal of Infectious Diseases 221(11):1770–1774.

Wang, L.; Lin, Z. Q.; and Wong, A. 2020. Covid-net: A tailored deep convolutional neural network design for detection of covid-19 cases from chest radiography images. arXiv preprint 2003.09871.

Wang, J.; Zhou, M.; and Liu, F. 2020. Reasons for healthcare workers becoming infected with novel coronavirus disease 2019 (covid-19) in china. Journal of Hospital Infection 105(1):100–101.

Wong, H. Y. F.; Lam, H. Y. S.; Fong, A. H.-T.; Leung, S. T.; Chin, T. W.-Y.; Lo, C. S. Y.; Lui, M. M.-S.; Lee, J. C. Y.; Chiu, K. W.-H.; Chung, T.; et al. 2020. Frequency and distribution of chest radiographic findings in covid-19 positive patients. Radiology 201160.

World Health Organization. 2020. Laboratory testing for 2019 novel coronavirus (2019-ncov) in suspected human cases, interim guidance, 2 march 2020.

Wu, Z.; and McGoogan, J. M. 2020. Characteristics of and important lessons from the coronavirus disease 2019 (covid-19) outbreak in china: summary of a report of 72 314 cases from the chinese center for disease control and prevention. Jama 323(13):1239–1242.

Xu, X.; Jiang, X.; Ma, C.; Du, P.; Li, X.; Lv, S.; Yu, L.; Chen, Y.; Su, J.; Lang, G.; et al. 2020. Deep learning system to screen coronavirus disease 2019 pneumonia. arXiv preprint 2002.09334.

Yadav, S. S.; and Jadhav, S. M. 2019. Deep convolutional neural network based medical image classification for disease diagnosis. Journal of Big Data 6(1):113.

Yang, P.; Ding, Y.; Xu, Z.; Pu, R.; Li, P.; Yan, J.; Liu, J.; Meng, F.; Huang, L.; Shi, L.; et al. 2020. Epidemiological and clinical features of covid-19 patients with and without pneumonia in beijing, china. Medrxiv.

Zhang, J.; Xie, Y.; Li, Y.; Shen, C.; and Xia, Y. 2020. Covid-19 screening on chest x-ray images using deep learning based anomaly detection. arXiv preprint 2003.12338.

Zhao, W.; Zhong, Z.; Xie, X.; Yu, Q.; and Liu, J. 2020. Relation between chest ct findings and clinical conditions of coronavirus disease (covid-19) pneumonia: a multicenter study. American Journal of Roentgenology 214(5):1072–1077.

Zhou, S.; Wang, Y.; Zhu, T.; and Xia, L. 2020. Ct features of coronavirus disease 2019 (covid-19) pneumonia in 62 patients in wuhan, china. American Journal of Roentgenology 1–8.

Zu, Z. Y.; Jiang, M. D.; Xu, P. P.; Chen, W.; Ni, Q. Q.; Lu, G. M.; and Zhang, L. J. 2020. Coronavirus disease 2019 (covid-19): a perspective from china. Radiology 200490.

